# Short communications of a new circulating Dengue genotype III American II lineage in Mexico: outbreak 2022-2023

**DOI:** 10.1101/2024.02.24.24303299

**Authors:** RL Madera-Sandoval, AP Rodríguez-Maldonado, SV Rivero-Arredondo, J Quiroz-Mercado, IE Molina-Gomez, L Hernandez-Rivas, I Lopez-Martinez, M Vazquez-Pichardo, F Correa-Morales, C Wong-Arámbula

**Affiliations:** Departamento de Biología Molecular y Validación de Técnicas, Instituto de Diagnóstico y Referencia Epidemiológicos “Dr, Manuel Martínez Báez” (InDRE), Secretaria de Salud, Mexico City, Mexico; Dirección de Servicios y Apoyo Técnico, Instituto de Diagnóstico y Referencia Epidemiológicos “Dr, Manuel Martínez Báez” (InDRE), Secretaria de Salud, Mexico City, Mexico; Dirección de Diagnóstico y Referencia, Instituto de Diagnóstico y Referencia Epidemiológicos “Dr, Manuel Martínez Báez” (InDRE), Secretaria de Salud, Mexico City, Mexico; Departamento de Virología, Instituto de Diagnóstico y Referencia Epidemiológicos “Dr, Manuel Martínez Báez” (InDRE), Secretaria de Salud, Mexico City, Mexico; Director de Vectores en el Centro Nacional de Programas Preventivos y Control de Enfermedades, CENAPRECE, México

**Keywords:** Dengue virus, Mexico, Americano II, genotype, Next-generation sequencing, Bioinformatics

## Abstract

We report for the first time the circulation of American II lineage of Dengue virus serotype 3 genotype III in Mexico uncovered by the genomic surveillance in charge of the Instituto de Diagnóstico y Referencia Epidemiológicos through sequencing the whole genome using Illumina platform. The importance of the characterization of a new lineage circulating in Mexico impacts public health and the actions to be taken to mitigate new outbreaks and in the future associate them with clinical responses that the new lineage may cause.

---

Major dengue (DENV) outbreaks have been identified in Mexico since the introduction of virus to the present, the serotypes can be divided in four (DENV-1; DENV-2; DENV-3; DENV-4) and various genotypes has been recorded (Domínguez-de-la-Cruz et al., 2020; Hernández-García et al., 2020). In 2022 and 2023 increase in cases of DENV has registered, 3,111 cases of DENV-1; 6,242 cases of DENV-2; 8,250 cases of DENV-3 and 348 cases of DNV-4 (Epidemiol, 2023); as a consequence of this increase in DENV cases an analysis of the genomic diversity of circulating viruses was carried out during this period and the genotypic diversity of each of the circulating serotypes was determined. Further, different reports by the countries of the Americas region (South America, Central America and the Caribbean) informed the introduction of new genotypes and linages of DENV-2 and DENV-3 (González M., 2023; Morel et al., 2023; Naveca et al., 2023; Paquita García et al., 2022).

One hundred forty samples characterized as DENV-3 by qPCR were extracted and sequenced using the NextSeq 500 Illumina platform, reads were mapped to a reference genome from the NCBI database for DENV-3 (DENV3_NC_001474) (DENV-3 can be divided into 4 genotypes I; II; III; V). A phylogenetic analysis was performed, showed a grouping of the set with genotype III (Fig. 1) which has been in circulation for the last few years and since the first outbreak by DENV-3 in 1996 (Domínguez-de-la-Cruz et al., 2020).

**Figure 1.**
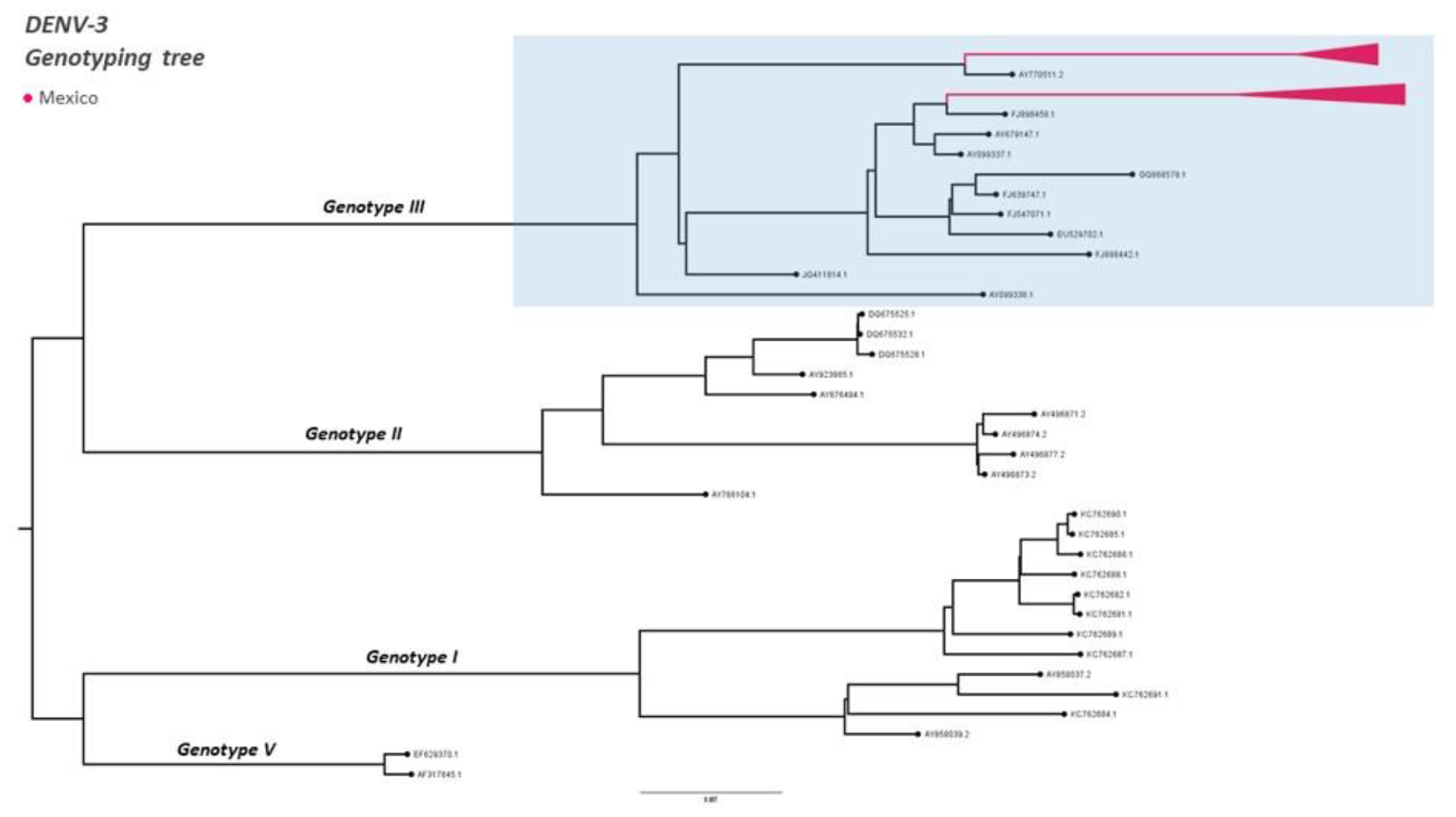
Identification of DENV-3 genotype III. A multiple sequence alignment was constructed with the MAFFT alignment software (Katoh and Standley, 2013)], using a subset by (Fonseca et al., 2019). The alignment was edited manually. The phylogenetic analysis inferred using IQ-TREE (Nguyen et al., 2015) (i.e. Maximum likelihood, 1000 bootstrap replicates)

Therefore, an analysis was carried out within the results obtained for genotype III which are divided in three lineages: Asian, American I and the last lineage detected in the Americas during 2022-2023 American II (Naveca F., 2023). The samples sequenced from 2022 and 2023 were found to belong to the American I lineage where they generally cluster. However, some samples of April and May 2023 started to be grouped in an American II lineage (Fig. 2), this is the first time that the circulation of this lineage in Mexico has been reported.

**Figure 2.**
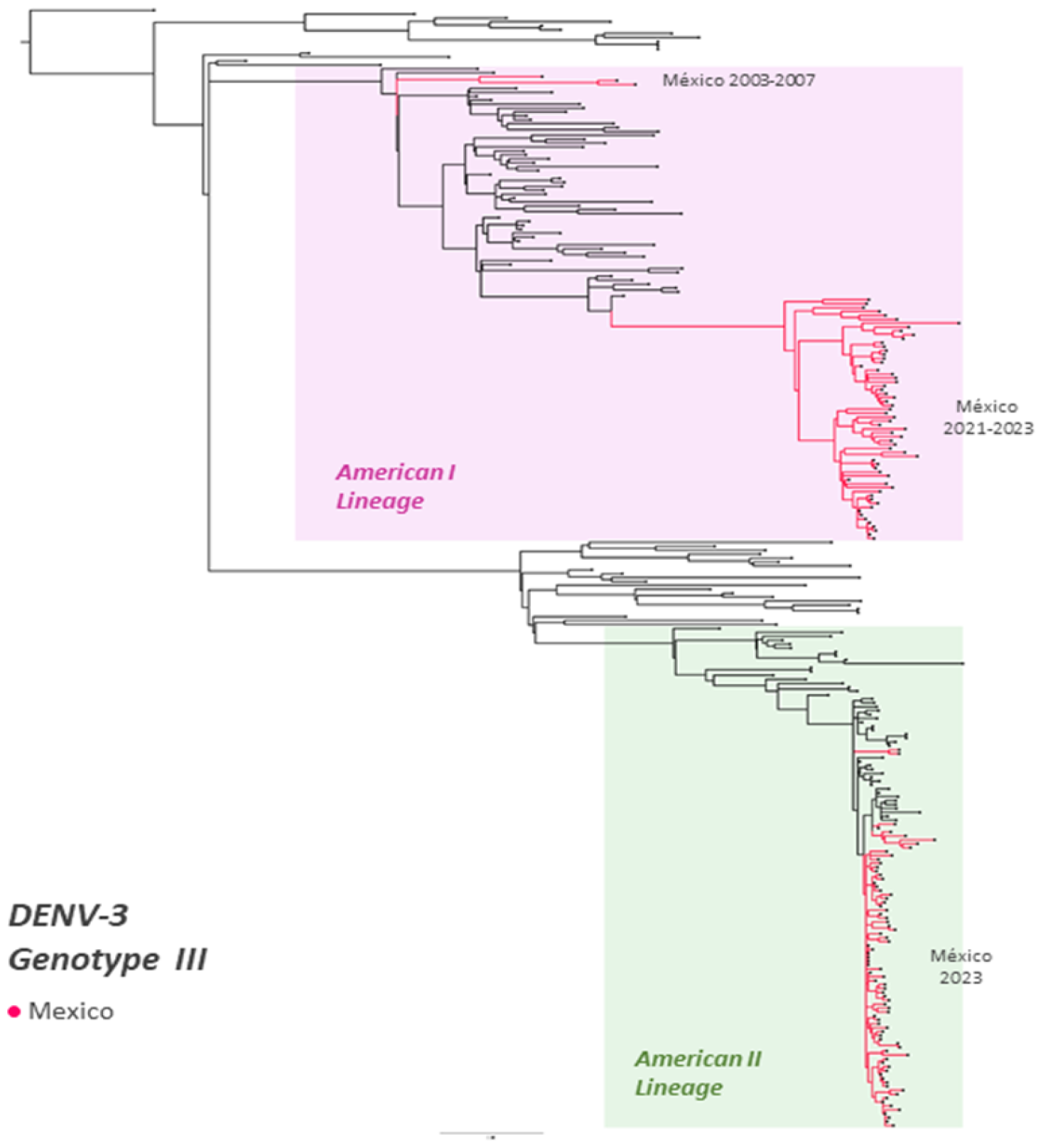
Identification of DENV-3, genotype III lineages. A multiple sequence alignment was constructed with the MAFFT alignment software [Katoh K, Standley DM. MAFFT multiple sequence alignment software version 7], using a subset by Santiago et al. 2023, unpublished. The alignment was edited manually. The phylogenetic analysis inferred using IQ-TREE ((Nguyen et al., 2015)) (i.e. Maximum likelihood, 1000 bootstrap replicates). *Branches outside the delimited American lineages belongs to Asian lineages.

The geographical distribution (Fig. 3) show that American II linage it is preserved in the central (Morelos, 10 samples; Queretaro, 1 sample; Jalisco, 1 sample;) and southern region of the country (Yucatan, 9 samples; Quintana Roo, 26 samples; Oaxaca, 1 sample; Chiapas, 5 sample and Veracruz, 35 sample), and American I linage circulates throughout the country Veracruz (8); Yucatan (10); Coahuila (3); Tabasco(8); Tamaulipas(1); Nuevo Leon(2); Puebla(18); Oaxaca(1); Chiapas (4); Hidalgo (1); Sonora (1); Quintano Roo (2) and Morelos (1). The introduction of this new circulating linage in Mexico its complicated to establishment, owing to, in the last 5 years there are no reported sequences in the repositories for neighboring countries to Mexico.

**Figure 3.**
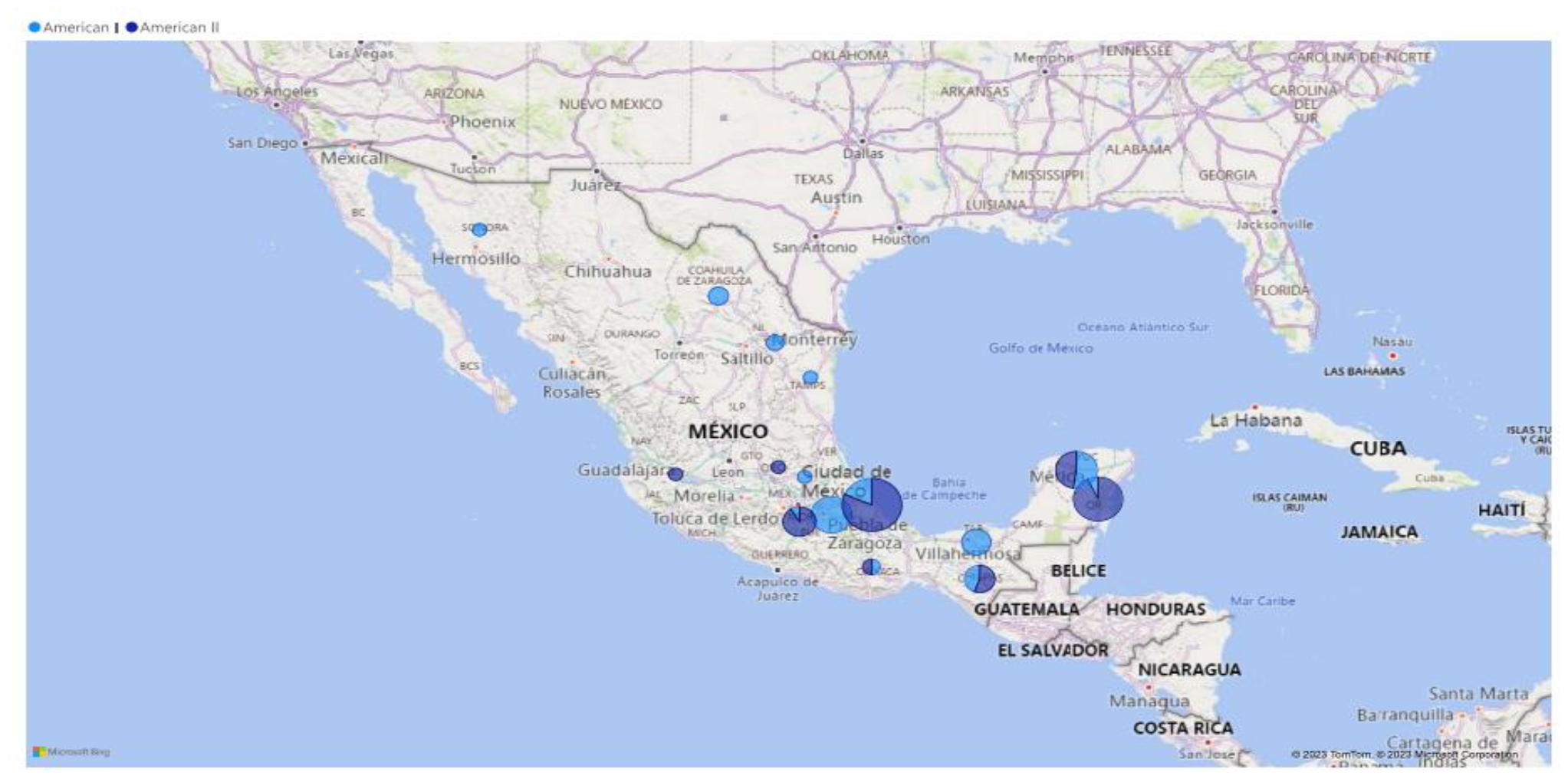
Geographic distribution by state of Mexico of serotype III, genotype III 2022-2023. Map of Mexico show the distribution of lineage American I (circle light blue) through the country and American II (circle dark blue) in the center and south of the country. Circles represents the proportions of samples for each linage.

This communication confirms the circulation of DENV-3, GIII-American II lineage in Mexico from 2023, and the expansion of this lineage is rapid, since the dates of the samples begin in the south and end in the center of Mexico. Our phylogenetic analysis cannot reveal where this lineage was introduced caused by lack of information from neighboring countries. However, our complete genome sequences will help to continue completing the phylogeny analysis of the different genotypes and their lineages of other interested parties.

The identification of a new lineage introduced into a susceptible population has direct consequences on public health, the molecular surveillance is valuable to monitoring rapid spread, circulation, and localized outbreaks (Do et al., 2023; Morel et al., 2023) of DENV-3-GIII American II lineage across the country’s territory and determinate the potential impact of this lineage in the population and to be able to relate them with the appearance of severe symptoms of the disease.

## Data Availability

All data produced in the present study are available upon reasonable request to the authors

## Ethical approval

The present study includes printed information; the samples used in this research were identified as a residuals samples from the routine diagnosis of arboviruses laboratory at the Instituto de Diagnóstico y Referencia Epidemiológicos.

## Author statement

WAC and RMAP conceptualized the study. RLMS wrote the manuscript. RLMS, RASV, QMJ performed all sequencing experiments, RMAP performed the bioinformatics and phylogenetic analysis and MGIE, HRL, LMI, VPM AND CMF reviewed the manuscript.

## Acknowledgments

To the Sistema Nacional de Vigilancia Epidemiológica de Enfermedades Transmitidas por Vector and a special mention for the Red Nacional de Laboratorios de Salud Pública de México for their valuable support with Epidemiological Laboratory Surveillance in each State of the Country.

To Dr. Jairo Mendez Rico and Dr. Leticia Franco of the Panamerican Health Organization, for the advice provided.

